# Multimodal MRI examination of structural and functional brain changes in older women with breast cancer in the first year of antiestrogen hormonal therapy

**DOI:** 10.1101/2022.02.25.22271510

**Authors:** Brenna C. McDonald, Kathleen M. Van Dyk, Rachael L. Deardorff, Jessica N. Bailey, Wanting Zhai, Judith E. Carroll, James C. Root, Tim A. Ahles, Jeanne S. Mandelblatt, Andrew J. Saykin

**Author notes:** Corresponding author: Brenna McDonald, PsyD. Co-First Authors (Brenna C. McDonald, Kathleen M. Van Dyk). Co-Senior Authors (Jeanne S. Mandelblatt, Andrew J. Saykin). ClinicalTrials.gov Identifier: NCT03451383. Data from this study were presented in part at the meeting of the International Cognition and Cancer Task Force, Denver, CO, February 2020.

## Abstract

**Purpose:** Cancer patients are concerned about treatment-related cognitive problems. We examined effects of antiestrogen hormonal therapy on brain imaging metrics in older women with breast cancer.

**Methods:** Women aged 60+ treated with hormonal therapy only and matched non-cancer controls (n=29/group) completed MRI and objective and self-reported cognitive assessment at pre-treatment/enrollment and 12 months later. Gray matter was examined using voxel-based morphometry (VBM), FreeSurfer, and brain age calculations. Functional MRI (fMRI) assessed working memory-related activation. Analyses examined cross-sectional and longitudinal differences and tested associations between brain metrics, cognition, and days on hormonal therapy.

**Results:** The cancer group showed regional reductions over 12 months in frontal, temporal, and parietal gray matter on VBM, reduced FreeSurfer cortical thickness in prefrontal, parietal, and insular regions, and increased working memory-related fMRI activation in frontal, cingulate, and visual association cortex. Controls showed only reductions in fusiform gyrus on VBM and FreeSurfer temporal and parietal cortex thickness. Women with breast cancer showed higher estimated brain age and lower regional gray matter volume than controls at both timepoints. The cancer group showed a trend toward decreased performance in attention, processing speed, and executive function over time. There were no significant associations between brain imaging metrics and cognition or days on hormonal therapy.

**Conclusion:** Older women with breast cancer showed brain changes in the first year of hormonal therapy. Increased brain activation during working memory processing may be a sign of functional compensation for treatment-related structural changes. This hypothesis should be tested in larger samples over longer time periods.

## Introduction

Breast cancer is the second most common cancer in US women, and over half of those diagnosed annually are ages 60 and older [1]. Seventy-five percent will be treated with 5-10 years of antiestrogen hormonal therapy (HT) [2–4], which is effective at reducing breast cancer recurrence through downregulating the action of estrogen in the body [3]. Estrogen is closely linked with cognitive function, especially in older women [5], and HTs cross the blood-brain barrier [6, 7], raising questions about the potential for cognitive side effects of this common treatment modality [6–8].

Despite years of clinical use, the extent to which HT affects cognition remains unclear due to limited research, inconsistent results, and differences in study design [9–21]. Recent results from the TAILORx trial comparing chemotherapy plus HT to HT alone showed self-reported cognitive concerns in both groups [22], and others have described worse self-reported cognition in women with breast cancer taking HT than non-cancer controls, despite little difference on cognitive testing [23]. We have also reported greater cognitive and other symptom burden in women with breast cancer on HT than non-cancer controls, and found that perceived cognitive difficulties are a risk factor for discontinuing HT [24, 25].

While extensively used to examine structural and functional brain changes after breast cancer chemotherapy [26–30], neuroimaging has not been widely applied to examine the impact of HT. Lower hippocampal volume and resting hippocampal-prefrontal functional connectivity have been reported in women with breast cancer taking tamoxifen [31, 32], and Hurria and colleagues found metabolic changes on FDG-PET in post-menopausal women with breast cancer taking an aromatase inhibitor, notably in regions relevant to aging [33]. HT exposure intersects with known changes to the brain and cognitive function associated with aging and menopause that are being actively investigated using neuroimaging. Therefore, better understanding of HT effects on the brain is clinically relevant for older women with breast cancer.

This single-site study examined a subset of the larger Thinking and Living with Cancer (TLC) prospective cohort [34, 35] who underwent neuroimaging. We used structural and functional MRI metrics to compare older women with breast cancer from pre-HT baseline to 12-month follow-up with matched non-cancer controls. We tested whether older women with breast cancer on HT exhibited different patterns of change in brain structure and function in the first year of therapy relative to controls, and explored if changes were related to self-reported and objective cognitive outcomes. Given the small number of participants available from this single-site sample, these analyses are intended to be hypothesis-generating to guide future research.

## Participants and Methods

The TLC Study is a national multicenter prospective cohort study (NCT03451383); overall study methods have been detailed previously [34, 35]. All data for this report were gathered between 2016-2020 at Indiana University (IU), the only TLC site conducting neuroimaging. Written informed consent was obtained from all participants according to the Declaration of Helsinki under a protocol approved by the IU Institutional Review Board.

### Participants

Women with breast cancer are eligible for the TLC study if they have a new diagnosis of primary non-metastatic breast cancer. Those with a history of other cancers (except non-melanoma skin cancer) are excluded if active treatment was <5 years or included systemic therapy. All participants (cancer and non-cancer control groups) are women aged ≥60 years; exclusion criteria include neurological or major psychiatric disorders or sensory impairment precluding assessment. All TLC participants at IU are offered the opportunity to participate in MRI scanning, and screened for MRI safety/claustrophobia if interested. For this study of effects of HT, we included women with breast cancer treated with adjuvant HT but not chemotherapy who had MRI data at both pre-HT baseline and 12-month follow-up (n=29). Two additional women with breast cancer who took HT for <2 months and one who was prescribed raloxifene primarily for osteoporosis were excluded from analysis. We selected 29 controls with baseline and 12-month follow-up data who best matched the cancer group for age and education as a comparison group.

### Data Collection

Assessments were completed at baseline (post-surgery, pre-radiation and/or HT) and 12-month follow-up, and included a survey capturing demographic, health history, and symptom information, neuropsychological testing, and neuroimaging. Clinical information was abstracted from medical records of women with breast cancer.

### Cognitive Measures

Objective cognitive assessment included domain-specific z-scores for attention, processing speed, and executive functioning (APE) and verbal learning and memory (LM). The APE domain consisted of the Digits Forward and Digits Backward subtests from the Neuropsychological Assessment Battery (NAB), Trail Making Tests A and B, the Controlled Oral Word Association Test, and the Digit Symbol subtest from the Wechsler Adult Intelligence Scale-III [36–39]. The LM domain consisted of the Logical Memory I and II subtests from the Wechsler Memory Scale-III and the Immediate Recall, Short Delayed Recall, and Long Delayed Recall scores from the NAB List Learning Test [38, 40]. Raw neuropsychological test scores were standardized to the baseline mean and standard deviation of the overall TLC control group, stratified by age and education, which were used to calculate the APE and LM domain scores.

Self-reported cognition was assessed using the Perceived Cognitive Impairments (PCI) score from the Functional Assessment of Cancer Therapy-Cognitive Function (FACT-Cog) [41].

### Psychosocial and Clinical Measures

We used several psychosocial and clinical measures to characterize the study sample. Baseline verbal intellect was estimated using the Wide Range Achievement Test-4 Word Reading subtest [42]. Depressive and anxiety symptoms were assessed with the Center for Epidemiologic Studies-Depression Scale (CES-D) and the State-Trait Anxiety Inventory (STAI) State subscale [43, 44]. Past use of menopausal hormone replacement therapy, age at menopause, and days on HT were based on self-report and data from the medical record.

### MRI scan acquisition

All scans were acquired on the same Siemens Prisma 3T scanner using a 64-channel head and neck coil. A T1-weighted three-dimensional magnetization prepared rapid gradient echo (MPRAGE) volume was used for voxel-based morphometry (VBM), FreeSurfer, and brain age calculations. A gradient-echo, echo-planar sequence was used for fMRI (see Online Resource 1 for sequence parameters).

### Working Memory fMRI Task

A visual-verbal n-back task was used to elicit brain activation during working memory processing, as in our previous studies [45–60] and as recommended by the International Cognition and Cancer Task Force [61] (see Online Resource 1 for details).

### Neuroimaging Preprocessing and Analysis

#### Voxel-Based Morphometry (VBM)

VBM was used to examine gray matter volume across the whole brain. T1-weighted images were processed using the standard longitudinal pipeline in Computational Anatomy Toolbox for SPM12 (CAT12.6). Statistical models were generated using a flexible factorial design to examine within- and between-group longitudinal changes, including group-by-time interactions, and a full factorial design to examine between-group cross-sectional differences (see Online Resource 1 for details). Overall significance was set at family-wise error (FWE) correction p=0.05. Clusters were considered significant at cluster-level p_uncorrected_<0.05. Participant mean values for clusters showing significantly reduced gray matter volume from baseline to follow-up were extracted using MATLAB for correlational analyses.

#### FreeSurfer

Cortical and volumetric segmentation was performed on T1-weighted images using the longitudinal FreeSurfer 6.0.0 pipeline [62]. A within-subject template image was created and used for skull stripping, Talairach transforms, and atlas registration prior to segmentation. For data reduction purposes, selected regional values were summed bilaterally to create prefrontal, temporal, parietal, basal ganglia, thalamus, limbic, and insular volumes and prefrontal, temporal, and parietal thicknesses (Online Resource 2). Participant mean values for regions showing significantly reduced gray matter thickness from baseline to follow-up were used for correlational analyses.

#### Brain Age

Predicted brain age values were generated with brainageR (version 1.0, https://github.com/james-cole/brainageR/releases/tag/1.0), which uses SPM12 and KernLab in R [63]. T1-weighted follow-up scans were registered to the baseline scan for each participant and then segmented. Predicted brain age was calculated using machine-learning Gaussian Process Regression (GPR) based on a training dataset of 2001 healthy individuals [64–66].

#### Task-Based fMRI

Preprocessing steps are detailed in Online Resource 1. Analyses focused on the most challenging working memory load condition (2-back>0-back). Statistical models were generated using a flexible factorial design to examine within- and between-group longitudinal changes, including group-by-time interactions, and a full factorial design to examine between-group cross-sectional differences. Overall cluster-level significance was set at p=0.001. Clusters were considered significant at cluster-level p_uncorrected_<0.05. Participant mean values for clusters showing significantly increased activation from baseline to follow-up were extracted using MATLAB for correlational analyses.

### Other Statistical Analyses

T-tests and chi-square tests were used for between-group comparisons of demographic, cognitive, psychosocial, and clinical variables, self-reported and objective cognition, and FreeSurfer and brain age variables, as well as between-group differences in change over time. For FreeSurfer volume variables, analyses controlled for baseline total intracranial volume. Within-group longitudinal comparisons used paired t-tests. In the cancer group, Pearson’s correlations examined relationships between change in neuroimaging variables, change in self-reported and objective cognitive outcomes, and days on HT. Analyses used SAS Version 9.4.b, and p<0.05 was considered statistically significant.

Given the minimal previous neuroimaging literature examining HT effects, we conducted three types of comparisons to best understand the data and generate hypotheses for future, larger studies. First, cross-sectional comparisons were conducted at each timepoint. It is important to examine baseline neuroimaging metrics in this older sample given pre-treatment neuroimaging differences found between younger women with breast cancer and controls [47, 67–70]. Cross-sectional differences at 12-month follow-up are also important for hypothesis generation, as our available sample size might limit power to detect differences in longitudinal change metrics. Second, within-group longitudinal change was examined to identify metrics showing significant alterations over 12 months. Third, we examined between-group differences in change over time (i.e., group-by-time interactions), to directly compare change in the cancer and control groups.

## Results

### Sample Characteristics

Groups were generally comparable at baseline (Table 1). The cancer group showed higher depressive symptoms (p<0.01) and a trend toward higher anxiety symptoms at baseline (p=0.06). Mean symptom levels were well below clinically meaningful thresholds [71, 72], however, and there were no significant between-group differences in change over time for depressive or anxiety symptoms (ps>0.30), so these variables were not included as covariates in analyses.

**Table 1.**
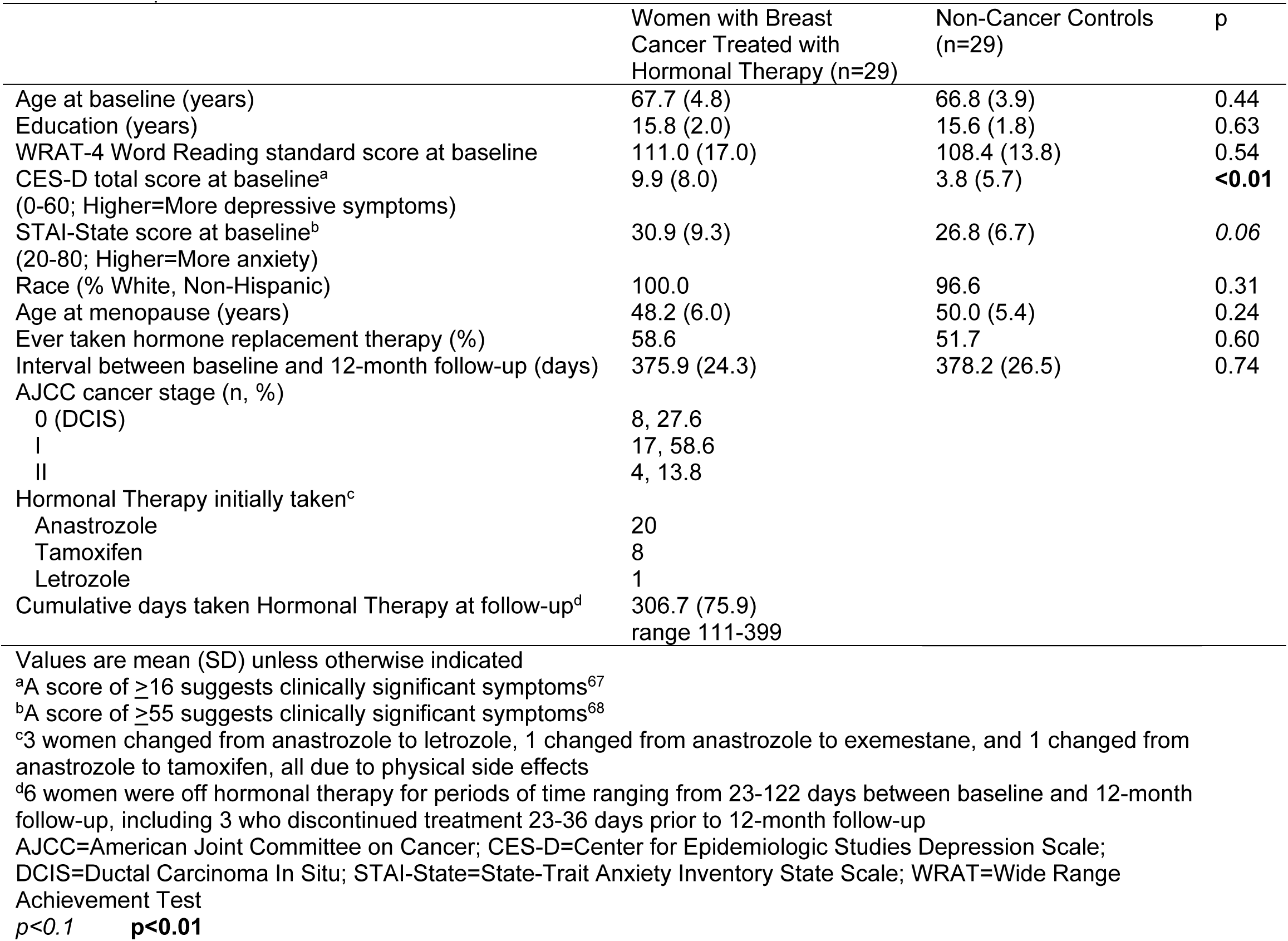
Sample Characteristics

### Objective and Self-Reported Cognitive Functioning

Self-reported and objective cognitive function did not differ between groups at baseline (Table 2). At follow-up the breast cancer group showed a trend toward lower performance than controls in APE (p=0.07). Between-group comparison of change scores showed a trend level difference (p=0.09) for this scale, with controls showing significantly improved performance over time (p=0.03), consistent with practice effects, while women with breast cancer showed no change. There were no other significant findings for self-reported or objective cognition.

**Table 2.**
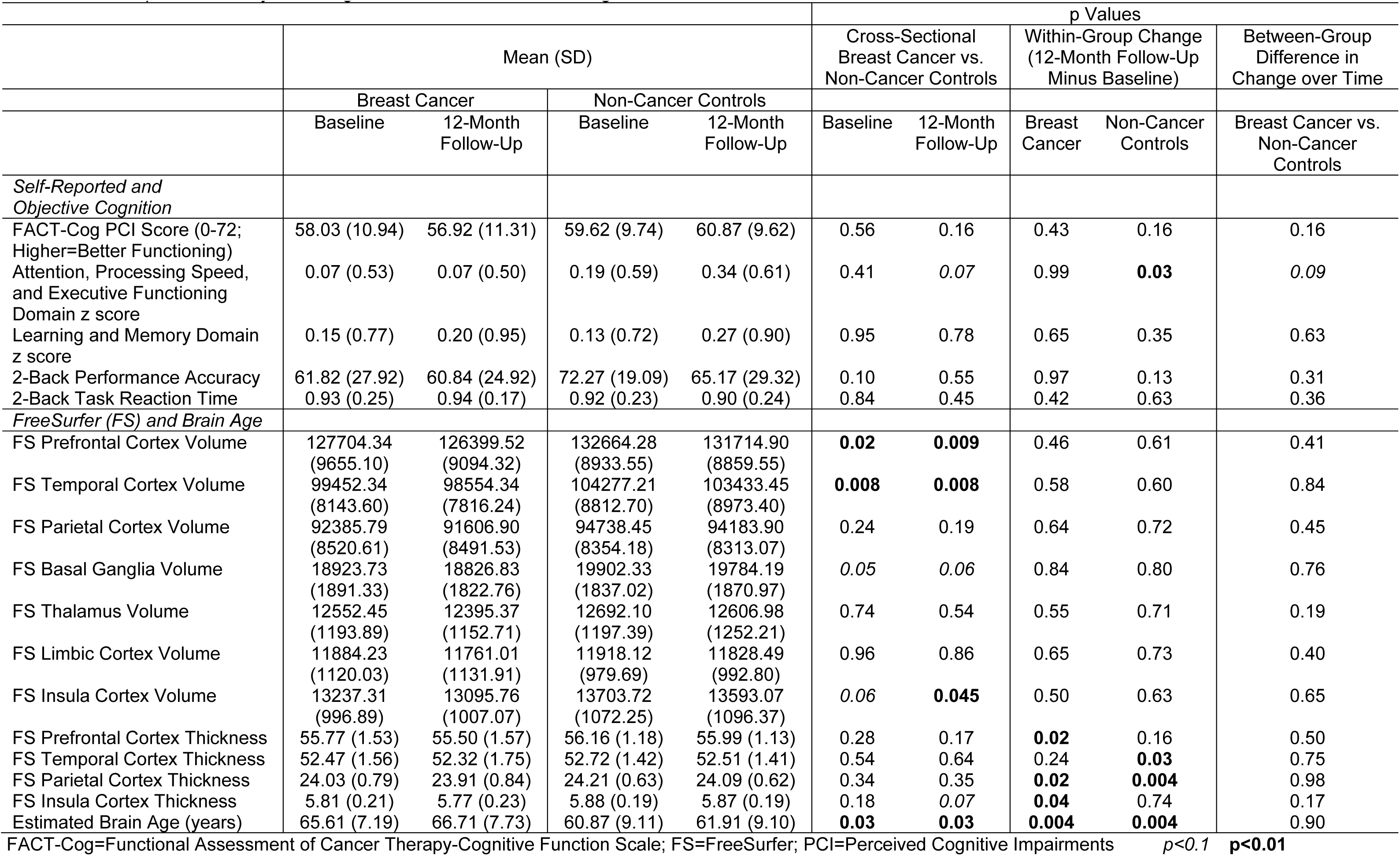
Self-Reported and Objective Cognition, FreeSurfer, and Brain Age Results

### Whole-Brain Voxel-Based Morphometry

Women with breast cancer showed significantly lower gray matter volume than controls in a right frontal cluster (BA6) at both baseline and follow-up (Figure 1, Table 3). The cancer group showed significantly reduced gray matter volume over time in left temporal (BA38) and parietal (BA40) and bilateral frontal regions (BA10, BA44, and BA47) (Figure 2A, Table 3). Controls showed a single cluster of significant gray matter volume reduction in the right fusiform gyrus (BA37) (Figure 2B, Table 3). There were no regions at either timepoint in which gray matter volume was significantly greater in women with breast cancer than controls, and no regions of increased volume over time in either group. No clusters were significant in group-by-time interactions.

**Fig. 1.**
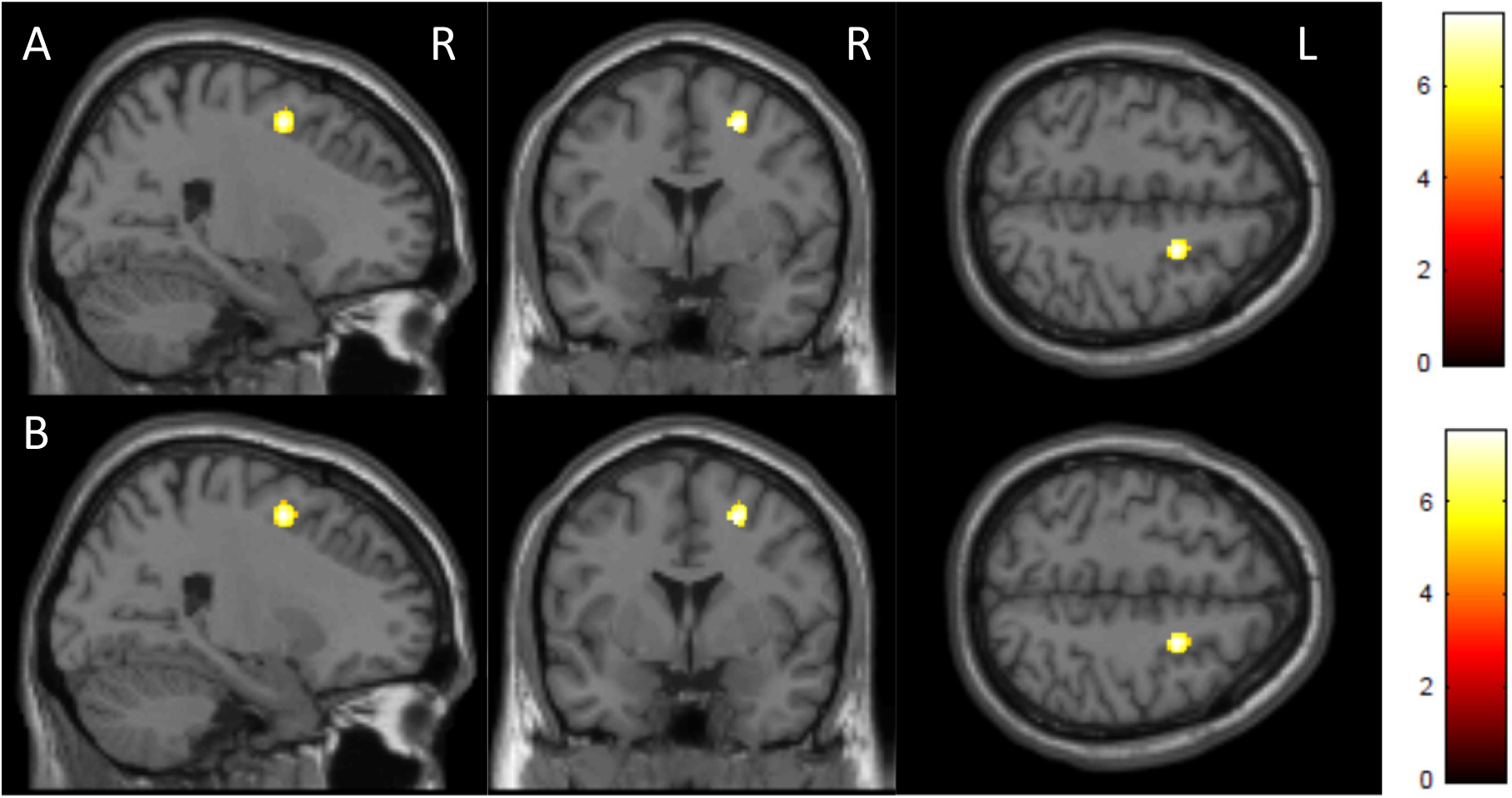
Voxel-based morphometry (VBM): Lower gray matter volume in older women with breast cancer treated with hormonal therapy relative to non-cancer controls at A) pre-treatment baseline and B) 12-month follow-up (Overall FWE-corrected p<0.05, cluster-level p_uncorrected_<0.05)

**Fig. 2.**
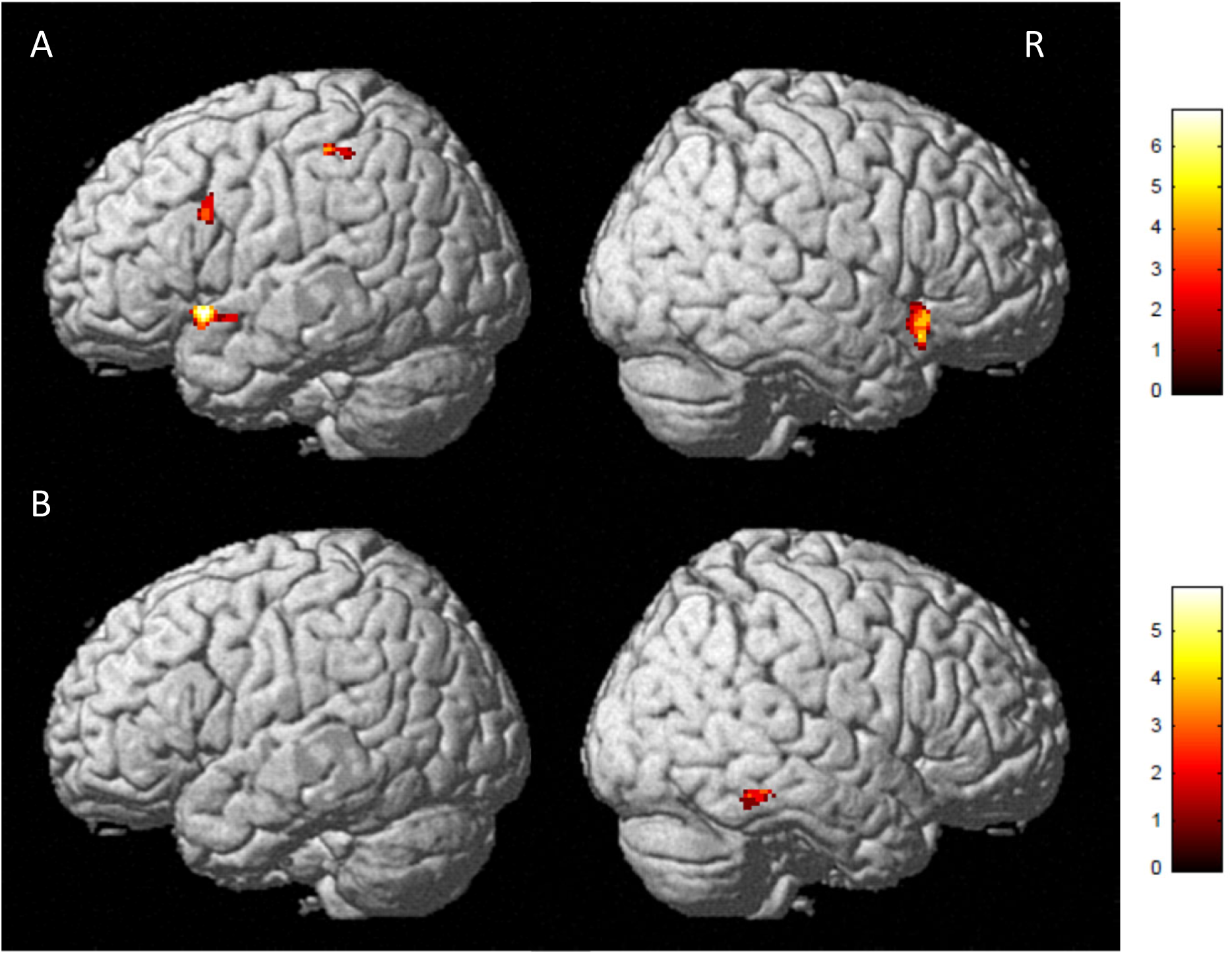
Voxel-based morphometry (VBM): Reduced gray matter volume from baseline to 12-month follow-up in A) Older women with breast cancer treated with hormonal therapy and B) Non-cancer controls (Overall FWE-corrected p<0.05, cluster-level p_uncorrected_<0.05)

**Table 3.** Voxel-Based Morphometry (VBM) Results (Overall FWE-corrected p<0.05, cluster-level p_uncorrected_<0.05)

| MNI Coordinates | k | Cluster-Level<br>$p_{\text{uncorrected}}$ | Z | Region Description |
| --- | --- | --- | --- | --- |
| Pre-Treatment Baseline: Non-Cancer Controls>Women with Breast Cancer Treated with Hormonal Therapy (Figure 1A) |  |  |  |  |
| 24 2 52 | 160 | 0.008 | 6.74 | Right premotor/supplementary motor cortex (BA6) |
| 12-Month Follow-Up: Non-Cancer Controls> Women with Breast Cancer Treated with Hormonal Therapy (Figure 1B) |  |  |  |  |
| 24 2 52 | 164 | 0.008 | 6.72 | Right premotor/supplementary motor cortex (BA6) |
| Pre-Treatment Baseline>12-Month Follow-Up: Women with Breast Cancer Treated with Hormonal Therapy (Figure 2A) |  |  |  |  |
| -51 14 -10 | 138 | 0.001 | 5.80 | Left temporal pole (BA38) |
| 44 20 -12 | 141 | 0.001 | 5.29 | Right pars orbitalis (BA47) |
| -52 12 30 | 47 | 0.033 | 5.24 | Left inferior frontal gyrus (BA44) |
| 0 51 15 | 55 | 0.023 | 5.02 | Right anterior prefrontal cortex (BA10) |
| -45 -40 51 | 39 | 0.049 | 4.92 | Left supramarginal gyrus (BA40) |
| Baseline>12-Month Follow-Up: Non-Cancer Controls (Figure 2B) |  |  |  |  |
| 64 -45 -18 | 43 | 0.040 | 5.14 | Right fusiform gyrus (BA37) |

BA=Brodmann Area; FWE=Family-Wise Error; k=cluster extent size (number of voxels); MNI=Montreal Neurological Institute

### FreeSurfer Regional Analyses

Women with breast cancer showed significantly lower volume than controls at both baseline and follow-up in prefrontal and temporal cortex (ps=0.02-0.008, Table 2). There was a trend for lower basal ganglia volume in the cancer group at both timepoints (ps=0.05-0.06). Insular volume showed a trend for lower volume in the cancer group at baseline, with increased significance at follow-up (ps=0.06 and 0.045, respectively). There were no significant within-group longitudinal changes in regional FreeSurfer volumes, nor between-group differences in change over time.

Women with breast cancer showed significant reduction over time in prefrontal, parietal, and insular cortex thickness (ps<0.05), as well as a trend toward lower insular cortex thickness relative to controls at follow-up (p=0.07). Controls showed reductions over time in temporal and parietal cortex thickness (ps=0.03-0.004). There were no significant between-group differences for cortical thickness at baseline or change over time.

### Brain Age

Estimated brain age for controls was ∼five years younger than chronological age on average, while estimates for women with breast cancer were comparable to chronological age; these significant group differences were observed at both baseline and follow-up (p=0.03, Table 2). There was no significant between-group difference in change in estimated brain age over time.

### Working Memory fMRI

There were no significant between-group differences in task-related activation at baseline, but at follow-up women with breast cancer showed significantly greater working memory-related activation than controls in right dorsolateral prefrontal (BA9) and dorsal posterior cingulate cortex (BA31) (Figure 3, Table 4). The cancer group showed significantly increased activation over time in right frontal (BA6, BA44, BA47) and visual association (BA19) and left frontal (BA6, BA9) and cingulate (BA23, BA32) cortex (Figure 4, Table 4). There were no regions in which women with breast cancer showed significantly decreased activation or in which controls showed significant change over time. No clusters were significant in group-by-time interactions.

**Fig. 3.**
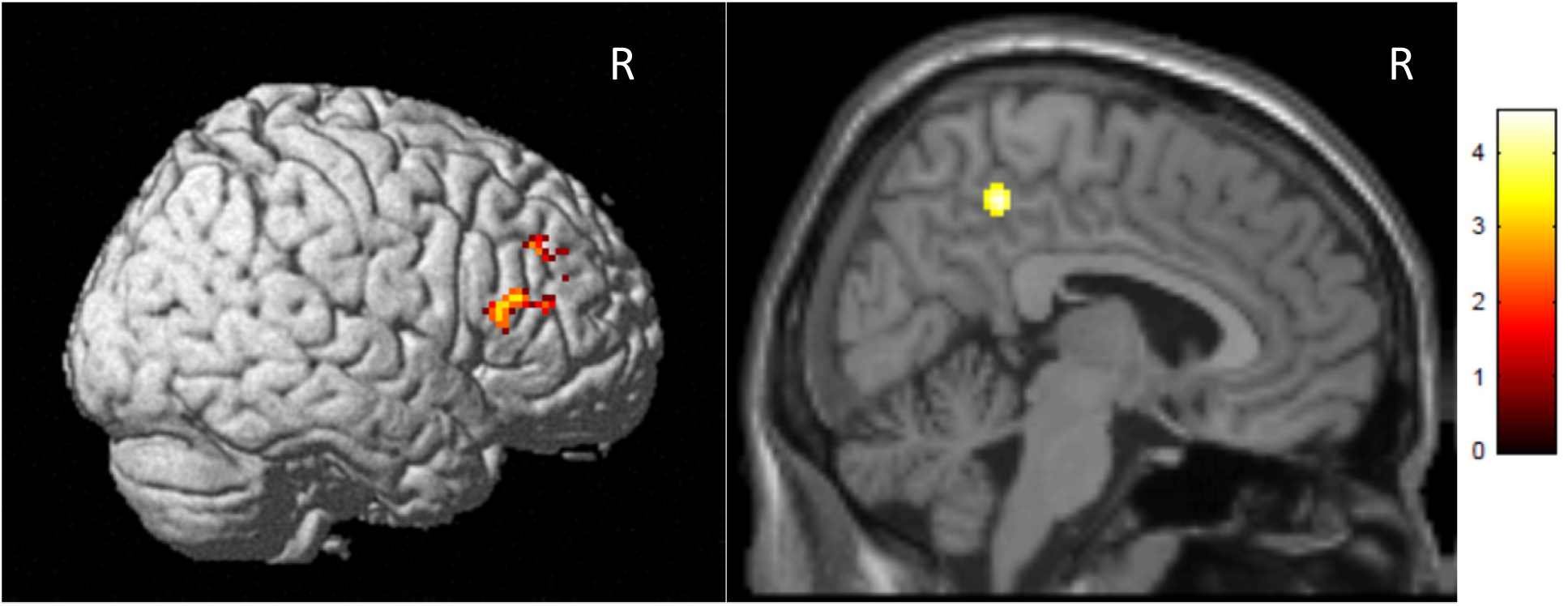
Greater working memory-related functional MRI activation (2-Back>0-Back) in older women with breast cancer treated with hormonal therapy relative to non-cancer controls at 12-month follow-up (Overall p<0.001, cluster-level p_uncorrected_<0.05)

**Fig. 4.**
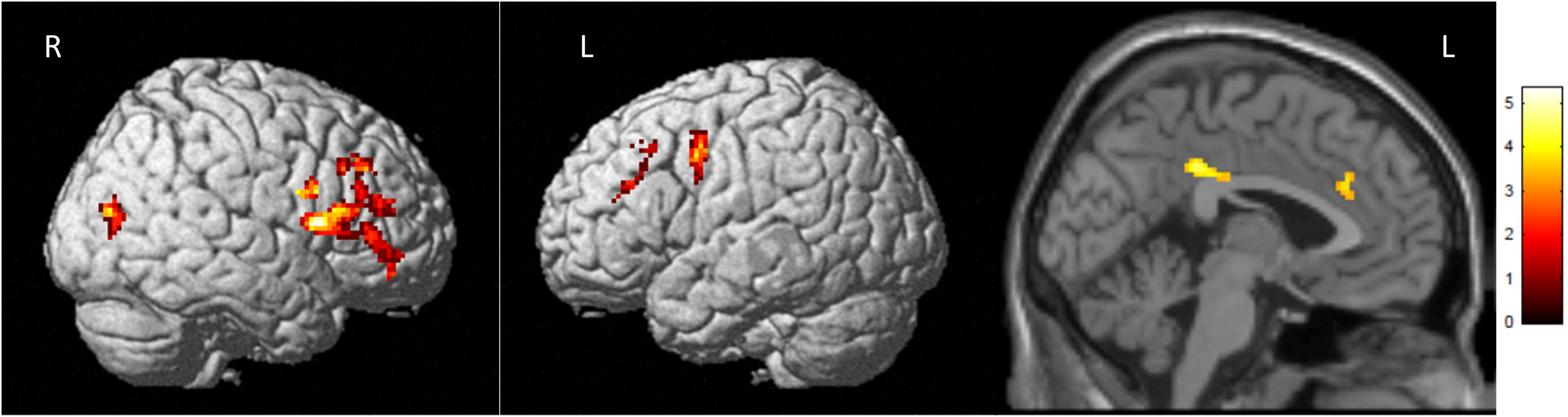
Increased working memory-related functional MRI activation (2-Back>0-Back) from baseline to 12-month follow-up in older women with breast cancer treated with hormonal therapy (Overall p<0.001, cluster-level p_uncorrected_<0.05)

**Table 4.** Working Memory-Related Functional MRI (fMRI) Results (overall p<0.001, cluster-level p_uncorrected_<0.05)

| MNI Coordinates | k | Cluster-Level<br>$p_{\text{uncorrected}}$ | Z | Region Description |
| --- | --- | --- | --- | --- |
| 12-Month Follow-Up: Women with Breast Cancer Treated with Hormonal Therapy > Non-Cancer Controls (Figure 3) |  |  |  |  |
| 22 42 32 | 107 | 0.025 | 4.33 | Right dorsolateral prefrontal cortex (BA9) |
| 6 -42 48 | 106 | 0.025 | 4.20 | Right dorsal posterior cingulate cortex (BA31) |
| 44 30 18 | 80 | 0.048 | 3.55 | Right dorsolateral prefrontal cortex (BA9) |
| 12-Month Follow-Up > Pre-Treatment Baseline: Women with Breast Cancer Treated with Hormonal Therapy (Figure 4) |  |  |  |  |
| -40 0 44 | 118 | 0.015 | 4.77 | Left premotor/supplementary motor cortex (BA6) |
| 52 14 10 | 453 | <0.001 | 4.62 | Right inferior frontal gyrus (BA44) |
| -8 30 26 | 164 | 0.005 | 4.61 | Left dorsal anterior cingulate cortex (BA32) |
| 38 4 26 | 113 | 0.016 | 4.35 | Right premotor/supplementary motor cortex (BA6) |
| -2 -36 36 | 92 | 0.028 | 4.05 | Left ventral posterior cingulate cortex (BA23) |
| 40 42 -2 | 93 | 0.027 | 3.93 | Right pars orbitalis (BA47) |
| 34 -76 18 | 78 | 0.040 | 3.79 | Right visual association cortex (BA19) |
| -36 32 26 | 87 | 0.032 | 3.67 | Left dorsolateral prefrontal cortex (BA9) |

BA=Brodmann Area; k=cluster extent size (number of voxels); MNI=Montreal Neurological Institute

### Relationships of Brain Changes to Cognition and Time on Treatment

There were no significant correlations between change in VBM gray matter volume, FreeSurfer thickness, or working memory-related activation in women with breast cancer and change in APE, LM, FACT-Cog PCI, or 2-back accuracy or reaction time (all ps>0.05). There were also no significant correlations between these variables and days on HT (all ps>0.05).

## Discussion

This is the first MRI study examining effects of the first 12 months of HT in older women with breast cancer. We used multiple structural and functional neuroimaging metrics and compared results to a demographically matched non-cancer control group. Women with breast cancer showed reductions in gray matter volume and cortical thickness over time that were not seen in controls. Brain age was higher in the cancer group than controls before therapy, an effect which persisted over time. While there were no significant associations between brain imaging metrics and cognition, women with breast cancer showed increased activation during working memory processing that was not seen in controls. Finally, in this first year of HT, there was no significant relationship between time on treatment and brain metrics. These results support the need for future examination of the role of HT in cancer-related cognitive decline over longer time periods.

Our findings in this small sample reveal a pattern of structural brain changes in the first year of HT. The few prior neuroimaging studies [31–33] of HT effects focused on single imaging modalities, single brain regions, or on women prescribed only tamoxifen or AIs; the one prior longitudinal study had only a six-month assessment interval. Given these limitations in comparability, it is noteworthy that the previous work also found regional gray matter reductions, as well as alterations in resting functional connectivity and cerebral metabolism related to HT. Our findings encourage the use of multiple, complementary imaging analytic methods and modalities in future work to increase sensitivity to detect differences in grey matter volume and thickness and brain activation.

Our finding that women with breast cancer had a higher estimated brain age and lower frontal and temporal gray matter volume than controls before systemic treatment is consistent with literature showing pre-treatment structural differences [67–70]. Such results are thought to reflect a potential contribution of other factors (e.g., other medical comorbidities, including cancer risk factors or disease processes, surgery or anesthesia exposure [73]) to brain function and cognition in individuals with cancer.

The observed neuroimaging changes were not directly associated with cognitive functioning or time on HT, raising several questions for future research. It may be that the first year of therapy is too limited an observation period (i.e., restricted range of length of time on HT). It is also possible that cumulative HT effects may take longer to manifest in detectable cognitive changes, or that the influence of HT is more complex than a function of additive exposure time. Notably, our results also suggest that functional compensatory mechanisms could be sustaining cognitive performance in the cancer group, despite the observed reductions in gray matter volume and thickness. We saw increased working memory-related brain activation in women with breast cancer over time, and greater activation in the cancer group than controls at follow-up. Given comparable task performance between groups, this may represent a compensatory response to maintain cognitive function, as has been hypothesized in other studies of women with breast cancer [45-47, 74, 75]. It will be important to examine larger, more heterogeneous samples and to follow women over a longer time period, given the potential for years of HT exposure. If increased brain activation is compensatory, this may suggest vulnerability for future cognitive changes when compensatory mechanisms and/or the benefits of higher baseline cognitive reserve are exhausted. HT exposure over the recommended 5-10 years has the potential to interact with other risks for cognitive decline (e.g., medical comorbidities, cognitive aging, dementia). Furthermore, while we only examined women who did not receive chemotherapy, HT and chemotherapy effects may be additive, especially as similar frontal and temporal regions appear vulnerable to effects of both types of treatment [26–30].

Much of the research studying cognitive effects of HT has focused on tamoxifen [8, 20, 21]. However, as in our study, post-menopausal women with breast cancer are most often prescribed aromatase inhibitors, which downregulate estrogen synthesis in the body and brain. Different classes of HT may have varying effects, but there were too few women on tamoxifen (only 28% of our sample) to compare regimen types. Future studies in broader age groups may be able to test hypotheses based on modes of action of different classes of HT. Overall, however, our findings suggest brain changes following HT exposure. These results are consistent with literature showing links between menopause, estrogen function, and changes in cognition and brain structure and function [76–80], as well as studies examining the potential for estrogen replacement therapy to protect the brain from cognitive decline and dementia [81, 82]. A prior fMRI study found greater working memory-related prefrontal activity in post-menopausal compared to pre- or peri-menopausal women [83], parallel to our finding that post-menopausal women with breast cancer on HT showed greater activation relative to controls. In a study of >500 post-menopausal women, younger brain age was associated with higher levels of circulating estrogen in APOE ε4 noncarriers, but the opposite pattern was seen in APOE ε4 carriers [84]. Since HTs cross the blood-brain barrier, their effects on brain estrogen levels are one biologically plausible explanation for brain changes and cognitive problems among women with breast cancer receiving HT.

This study used a comprehensive battery of MRI and cognitive metrics to generate clinically relevant hypotheses to fill gaps in knowledge about unanswered questions regarding effects of HT. Given the preliminary nature of our observations, there are several limitations that should be considered in evaluating the results and planning future studies. The sample size is small, which may have limited our ability to detect significant differences between groups or over time; however, our early data will be useful to inform power estimates for future studies. Clinical guidelines recommend that women diagnosed with hormone receptor positive breast cancers receive HT, so there were only four women with breast cancer in our neuroimaging study cohort who did not receive systemic treatment. Thus, we could not evaluate the effects of cancer itself over time separately from therapy. Participants were largely white; it is especially critical to study relationships between HT, cognition, and brain structure and function in minoritized individuals and samples from diverse backgrounds, who may have unique patterns of effects and contributing factors. Our sample was well-educated and likely representative of high cognitive reserve; it will be important to examine women with varying levels of cognitive reserve to discern potential differences in effects. The protocol also did not include testing of FSH/LH levels to explore effects of estrogen status. Finally, since this is an observational study, treatment type and adherence were subject to individual differences and introduced exposure variability. One opportunity to accelerate knowledge would be to add neuroimaging and cognitive assessments to randomized HT trials.

Overall, we found a clinically relevant pattern of structural and functional brain changes in post-menopausal women with breast cancer on HT that was not seen in non-cancer controls. HT is prescribed for the majority of women with breast cancer and is effective for preventing cancer recurrence. If confirmed, however, our results suggest there may be potential side effects from downregulating estrogen function in the brain among post-menopausal women. It will be important to replicate and extend our findings to longer-term outcomes in larger samples. Such studies will also be useful to determine which factors place women at greatest risk for brain changes on HT (e.g., APOE ε4 genotype [85] or comorbidities), since any adverse brain effects can have a broad impact on quality of life and health care costs among the growing population of older breast cancer survivors.

## Data Availability

The datasets generated and/or analyzed for the current study are not publicly available due to the ongoing nature of the study, but are available from the corresponding author on reasonable request.

## Acknowledgements

The authors gratefully thank all the participants who graciously gave their time and effort to this work, and our research MRI technologists and neuroradiologist colleagues for their efforts to ensure the successful completion of the study. We also thank Michiel B. de Ruiter, PhD, for his assistance with brain age calculations and analysis. Finally, we wish to posthumously acknowledge the contributions of John D. West, MS to this research.

## Statements & Declarations

### Author Contributions

All authors contributed to the study conception and design. Material preparation, data collection, and analysis were performed by Brenna McDonald, Rachael Deardorff, Jessica Bailey, and Wanting Zhai. The first draft of the manuscript was written by Brenna McDonald, Kathleen Van Dyk, and Jeanne Mandelblatt, and all authors commented on previous versions of the manuscript. All authors read and approved the final manuscript.

### Ethics Approval

This study was performed in line with the principles of the Declaration of Helsinki. Written informed consent was obtained from all participants under a protocol approved by the Indiana University Institutional Review Board (#1602851718, first approved July 27, 2016).

## Online Resource 1 Supplementary Methodological Information

### Scan Parameters

MPRAGE: repetition time (TR)=2300ms, echo time (TE)=2.95ms, flip angle (FA)=9°, field of view (FOV)=270×253mm, 256×240 matrix, 176 1.2mm-thick contiguous sagittal slices, voxel size 1.1×1.1×1.2mm, scan time 5:12

fMRI: TR=1200 ms, TE=29ms, FA=65°, FOV=220×220mm, 88×88 matrix, number of excitations (NEX)=1, multi-band=3, 54 interleaved 2.5mm-thick contiguous axial slices, voxel size 2.5×2.5×2.5mm, scan time 6:10

A 3D fluid-attenuated inversion recovery (FLAIR) sequence was also acquired, and reviewed along with the MPRAGE by a board-certified neuroradiologist to rule out incidental pathology. Image quality was assessed during scan acquisition and at multiple preprocessing steps (e.g., accuracy of skull stripping, alignment of individual images to templates).

### Working Memory fMRI Task

During scanning, participants saw a string of consonant letters (except L, W, and Y) presented one every three seconds. Task conditions were 0-, 1-, and 2-back, in a blocked design. For each consonant, participants used a button press device to signify whether the current letter was a match (i.e., was the same as the designated target or the letter presented 1- or 2-back in the sequence) or a nonmatch. Each condition was presented in 27-second epochs preceded by three seconds of instruction (e.g., “the match is one back”). The three experimental conditions were each presented three times in pseudorandom order for a total of nine task blocks. Participants rehearsed a practice version of the task before scanning to ensure that they understood the demands of the task. Stimuli were presented through an MRI-compatible projection system and programmed in Presentation, which recorded response accuracy and reaction times.

### Voxel-Based Morphometry (VBM) Preprocessing

VBM was used to examine gray matter volume across the whole brain using the standard longitudinal pipeline in Computational Anatomy Toolbox for SPM12 (CAT12.6). T1-weighted follow-up scans were registered to the baseline scan for each participant. Scans were then registered to the Montreal Neurological Institute (MNI) T1-weighted template and segmented into gray matter, white matter, and cerebrospinal fluid compartments using the MNI T1-weighted template and corresponding tissue probability maps. Gray matter maps were then spatially normalized to MNI space, resampled to 1.5mm isotropic voxels, and smoothed using an isotropic Gaussian spatial filter (FHWM=8mm) to reduce residual inter-individual variability. The smoothed, normalized gray matter maps were subjected to statistical parametric mapping on a voxel-by-voxel basis using the general linear model as implemented in SPM12 using total intracranial volume (generated by CAT12.6) as a covariate. The SPM prior probability gray matter template was used to restrict the statistical comparisons to the gray matter compartment. Statistical models were generated using a flexible factorial design to examine within- and between-group longitudinal changes, including group-by-time interactions (factors: 1) subject, baseline and follow-up scans for each participant, 2) group, two independent levels, breast cancer and control, 3) time, two non-independent levels, baseline and follow-up), and a full factorial design to examine between-group cross-sectional differences (factors: 1) group, two independent levels, breast cancer and control, 2) time, two non-independent levels, baseline and follow-up).

### Task-Based fMRI Preprocessing

For each participant’s echo-planar fMRI series the susceptibility-induced field was estimated and corrected using reverse phase-encoded imaging pairs with FSL 6.0.0 Topup [1]. Slice timing correction was performed with a slice order file using slicetimer in FSL. Corrected fMRI data were then processed with Multivariate Exploratory Linear Optimized Decomposition into Independent Components (MELODIC version 3.15) using the following parameters: high pass filter cutoff=100s, MCFLIRT motion correction, and spatial smoothing FWHM=6mm. Auto classification and removal of ICA components were completed with FMRIB’s ICA-based Xnoiseifier (FIX) version 1.06 [2, 3] using an in-house generated independent training set of 23 protocol-specific, age-matched healthy controls. Filtered data were then coregistered and normalized to MNI 2×2×2mm^3^ standard space and statistical parametric mapping on a voxel-by-voxel basis was conducted by using a general linear model approach using motion parameters as regressors in SPM12. Contrast images comparing task conditions were created for each participant for second-level multi-subject/between-group analyses. The SPM prior probability gray matter template was used to restrict the statistical comparisons to the gray matter compartment. Statistical models were generated using a flexible factorial design to examine within- and between-group longitudinal changes, including group-by-time interactions (factors: 1) subject, baseline and follow-up scans for each participant, 2) group, two independent levels, breast cancer and control, 3) time, two non-independent levels, baseline and follow-up), and a full factorial design to examine between-group cross-sectional differences (factors: 1) group, two independent levels, breast cancer and control, 2) time, two non-independent levels, baseline and follow-up).

## Online Resource 2 FreeSurfer Region Groupings by FreeSurfer Variable Name

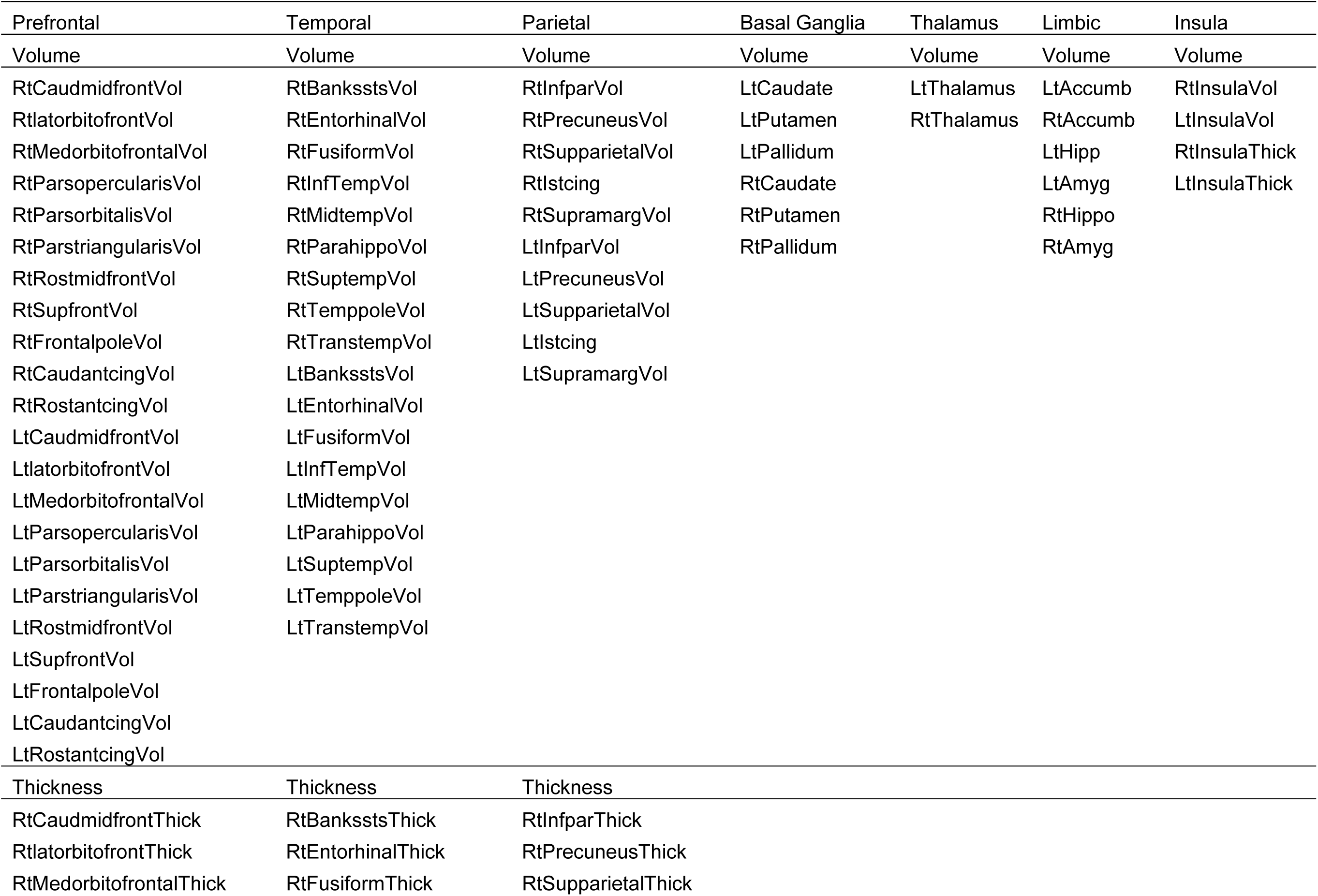
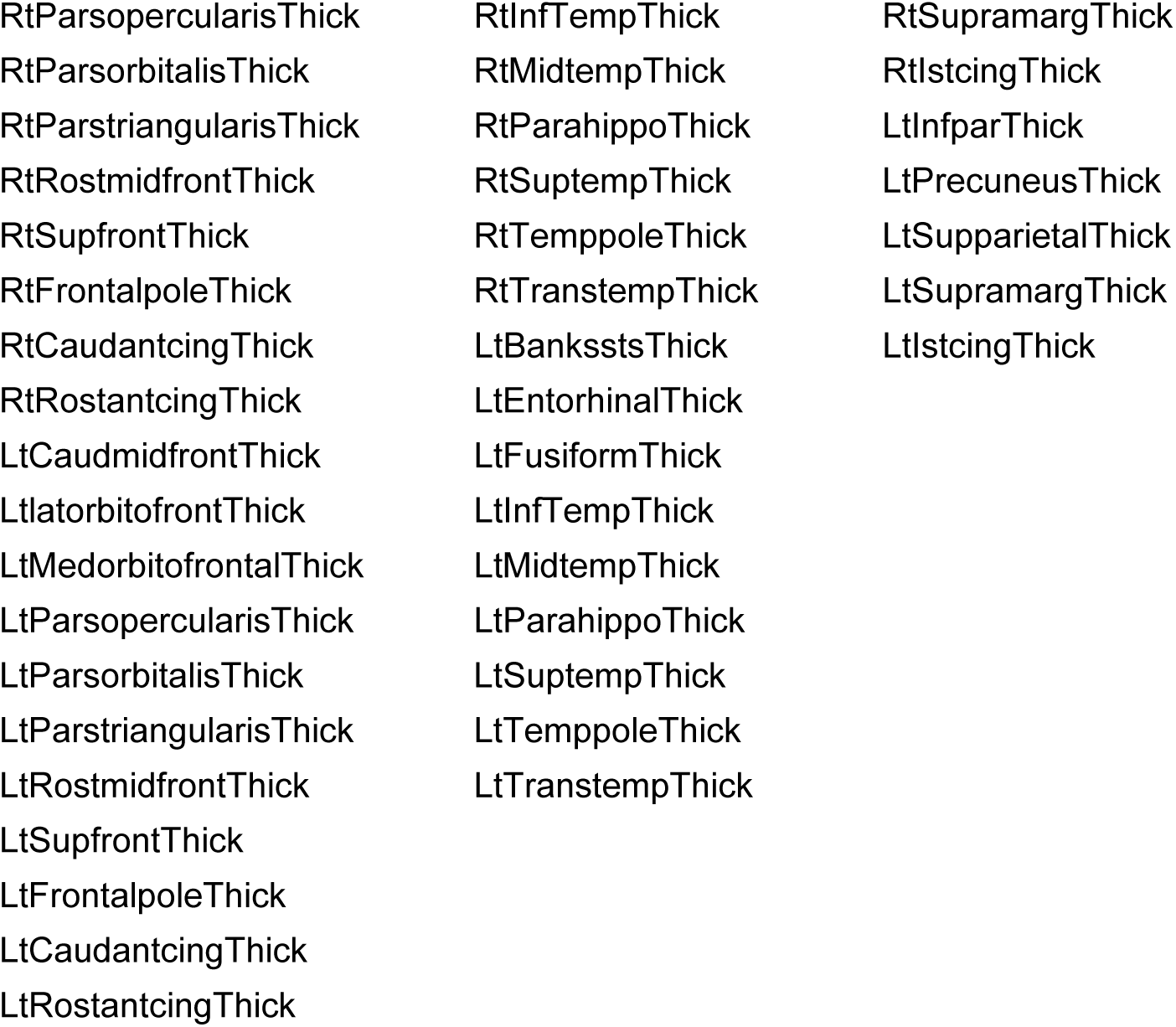

| Prefrontal | Temporal | Parietal | Basal Ganglia | Thalamus | Limbic | Insula |
| --- | --- | --- | --- | --- | --- | --- |
| Volume | Volume | Volume | Volume | Volume | Volume | Volume |
| RtCaudmidfrontVol | RtBanksstsVol | RtInfparVol | LtCaudate | LtThalamus | LtAccumb | RtInsulaVol |
| RtlatorbitofrontVol | RtEntorhinalVol | RtPrecuneusVol | LtPutamen | RtThalamus | RtAccumb | LtInsulaVol |
| RtMedorbitofrontalVol | RtFusiformVol | RtSupparietalVol | LtPallidum |  | LtHipp | RtInsulaThick |
| RtParsopercularisVol | RtInfTempVol | RtIstcing | RtCaudate |  | LtAmyg | LtInsulaThick |
| RtParsorbitalisVol | RtMidtempVol | RtSupramargVol | RtPutamen |  | RtHippo |  |
| RtParstriangularisVol | RtParahippoVol | LtInfparVol | RtPallidum |  | RtAmyg |  |
| RtRostmidfrontVol | RtSuptempVol | LtPrecuneusVol |  |  |  |  |
| RtSupfrontVol | RtTemppoleVol | LtSupparietalVol |  |  |  |  |
| RtFrontalpoleVol | RtTranstempVol | LtIstcing |  |  |  |  |
| RtCaudantcingVol | LtBanksstsVol | LtSupramargVol |  |  |  |  |
| RtRostantcingVol | LtEntorhinalVol |  |  |  |  |  |
| LtCaudmidfrontVol | LtFusiformVol |  |  |  |  |  |
| LtlatorbitofrontVol | LtInfTempVol |  |  |  |  |  |
| LtMedorbitofrontalVol | LtMidtempVol |  |  |  |  |  |
| LtParsopercularisVol | LtParahippoVol |  |  |  |  |  |
| LtParsorbitalisVol | LtSuptempVol |  |  |  |  |  |
| LtParstriangularisVol | LtTemppoleVol |  |  |  |  |  |
| LtRostmidfrontVol | LtTranstempVol |  |  |  |  |  |
| LtSupfrontVol |  |  |  |  |  |  |
| LtFrontalpoleVol |  |  |  |  |  |  |
| LtCaudantcingVol |  |  |  |  |  |  |
| LtRostantcingVol |  |  |  |  |  |  |
| Thickness | Thickness | Thickness |  |  |  |  |
| RtCaudmidfrontThick | RtBanksstsThick | RtInfparThick |  |  |  |  |
| RtlatorbitofrontThick | RtEntorhinalThick | RtPrecuneusThick |  |  |  |  |
| RtMedorbitofrontalThick | RtFusiformThick | RtSupparietalThick |  |  |  |  |

|  |  |  |
| --- | --- | --- |
| RtParsopercularisThick | RtInfTempThick | RtSupramargThick |
| RtParsorbitalisThick | RtMidtempThick | RtIstcingThick |
| RtParstriangularisThick | RtParahippoThick | LtInfparThick |
| RtRostmidfrontThick | RtSuptempThick | LtPrecuneusThick |
| RtSupfrontThick | RtTemppoleThick | LtSupparietalThick |
| RtFrontalpoleThick | RtTranstempThick | LtSupramargThick |
| RtCaudantcingThick | LtBanksstsThick | LtIstcingThick |
| RtRostantcingThick | LtEntorhinalThick |  |
| LtCaudmidfrontThick | LtFusiformThick |  |
| LtlatorbitofrontThick | LtInfTempThick |  |
| LtMedorbitofrontalThick | LtMidtempThick |  |
| LtParsopercularisThick | LtParahippoThick |  |
| LtParsorbitalisThick | LtSuptempThick |  |
| LtParstriangularisThick | LtTemppoleThick |  |
| LtRostmidfrontThick | LtTranstempThick |  |
| LtSupfrontThick |  |  |
| LtFrontalpoleThick |  |  |
| LtCaudantcingThick |  |  |
| LtRostantcingThick |  |  |

